# The relative effects of staying in school and attending university on smoking, BMI and systolic blood pressure: evidence from multivariable Mendelian randomization

**DOI:** 10.1101/2023.06.29.23292030

**Authors:** Eleanor Sanderson, Neil M. Davies

## Abstract

**Objectives:** To investigate which levels of educational attainment affect health.

**Design:** Multivariable Mendelian randomization study (MVMR).

**Setting:** UK Biobank.

**Participants:** European ancestry participants born in England.

**Exposure Educational:** attainment was defined as leaving school before age 18, leaving school after 18, or getting a university degree. Randomly allocated genetic variants were used as instruments for these traits.

**Main outcome measures:** Body mass index (BMI), smoking initiation, and systolic blood pressure.

**Results:** The MVMR estimates provided little evidence that remaining in school to age 18 affected BMI (mean difference=0.04, 95% CI: -0.42, 0.50), but evidence getting a degree reduced BMI by 0.47 standard deviation 95% CI: (0.01 to 0.97). The MVMR estimates provided evidence that remaining in school to age 18 reduced the odds of initiating smoking (odds ratio (OR): 0.48, 95% CI: 0.30 to 0.76), whereas it provided little evidence of effects of getting a degree (OR: 1.14, 95% CI: 0.69 to 1.88). MVMR suggested that both remaining in school to age 18 and getting a degree had similar effects on systolic blood pressure (mean difference=-2.60 95% CI: -3.73 to -1.46 and mean difference=-3.63 95% CI: -4.92 to -2.34, respectively).

**Conclusions:** Multivariable Mendelian randomization can be used to estimate the effects of complex longitudinal exposures such as educational attainment. This approach can help elucidate how and when factors such as educational attainment affect health outcomes.

**Key messages:**

1. Previous studies have estimated the average effect an additional year of education has on health, irrespective of educational level.
2. However, this assumes that each educational level has the same effect, e.g., an additional year of education in primary school has the same effect as an additional year at university; this assumption is implausible.
3. Multivariable Mendelian randomization can be used to relax this assumption and estimate the independent effects of educational levels.
4. Previously reported effects of education on smoking initiation appear to be due to remaining in school until age 18. In contrast, effects on BMI are due to attending university, and effects on systolic blood pressure are similar across education levels.

## Introduction

Educational attainment is associated with important health and socioeconomic outcomes across life and represents a critical modifiable risk factor for improving population health.^1-7^ Educational attainment is typically measured as a continuous variable indicating the number of years of education someone has. However, this continuous definition of education imposes strong assumptions on analyses, typically including that each additional year of education has the same effect or that the study is interested in a weighted average of the effect of all education levels. This is unlikely to be a plausible assumption. An additional year of education at age 16 is likely to affect different individuals and have different consequences than an additional year at age 20. Thus, the marginal effect of an extra year is likely to differ depending on each stage of education. In addition, policymakers and individuals are likely to be more interested in the effects of completing each stage of education.

The association of levels of educational attainment and later health outcomes are likely to provide unreliable evidence because unmeasured factors, such as sociodemographic characteristics or family background, confound the relationship between education and health outcomes. Researchers have sought to overcome this problem using two forms of natural experiments (instruments): policy reforms that affected the amount of education and genetic variants associated with educational attainment, an approach known as Mendelian randomization (MR). Studies using policy reforms identify the effects of a specific educational change, e.g. the raising of the school leaving in 1972 in the UK identifies the effect of an additional year of education at age 15 in those who would not remain in school without the reform.^1 8-10^ Whereas other natural experiments identify the effects of different educational outcomes (e.g. distance to university identifies the effect of getting a university degree on those choosing to attend or not is affected by their distance to university).^11 12^

Mendelian randomization uses genetic variants and instrumental variables for years of educational attainment to estimate the causal effect of the years of education.^13 14^ Mendelian randomization relies on three key assumptions. First, the instruments must strongly associate with the exposure; second, there must be no unmeasured confounders of the instruments-outcome association; and finally-there must be no effect of the instruments on the outcome that is not mediated via the exposure. As the genetic variants used as instruments in Mendelian randomization are fixed across an individual’s lifetime, Mendelian randomization estimates are often interpreted as ‘lifetime effects’.^13 15^ Specifically, Mendelian randomization estimates a weighted average of the effects of genetically predicted years of education across the entire life course.^16^ The individual effects of an additional year of education are likely to differ across the levels of education, so the overall effect can be interpreted as the average effect of an additional year of educational attainment weighted by the effects of the genetic variants on the likelihood of achieving each level of educational attainment.

An alternative assumption is to identify the effects on the outcome of binary variables for each stage of education (i.e. the effect of obtaining a specific level of education, e.g. primary school, secondary school, and university). Mendelian randomization can be used to estimate the effects of a single binary definition of education (e.g., the effects of getting a university degree on health). However, these estimates will also reflect the effects of the other educational levels required to attend university if there are shared genetic effects of the different educational stages. For example, suppose a variant associated with remaining in school to age 18 was also associated with getting a university degree. In that case, the Mendelian randomization estimate of a single exposure will reflect the effect of staying in school until age 18 and getting a university degree. This is illustrated in **Figure 1**, where a Mendelian randomization estimate of the effects of getting a degree will not reflect only the direct effect of getting a degree but also any direct effects of earlier educational outcomes on later health.

**Figure 1.**
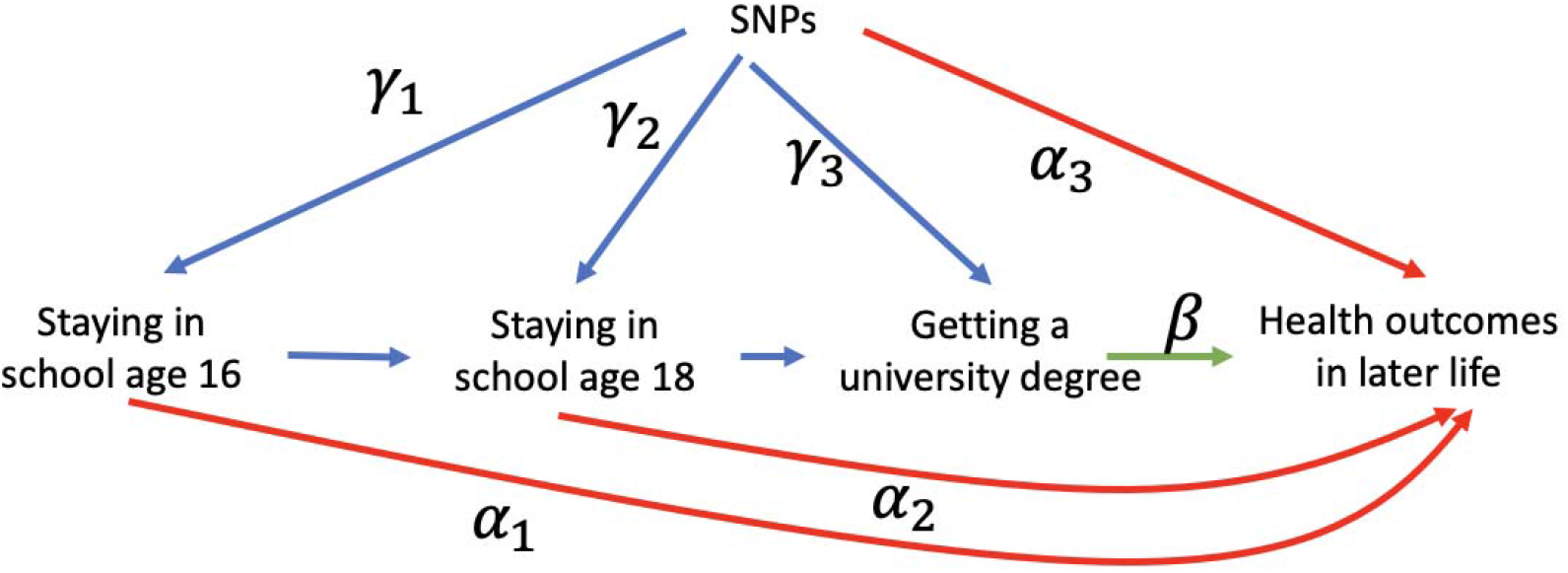
A directed acyclic graph of a Mendelian randomization estimation of the effects of remaining in school to ages 16 and 18 and getting a university degree on health. The effect of attending university on health outcomes (can be identified using multivariable Mendelian randomization by genetic variants affecting educational attainment under the following assumptions. First, the variants must explain sufficient variation in each level of educational attainment conditional on the other levels of educational attainment (the conditional relevance assumption). Second, there must be no direct effects of the SNPs on the outcome, except those mediated via educational attainment (). Thus, we can identify the effects of each educational level, and the pleiotropic effect due to an earlier educational level or initial cognitive ability will be accounted for (i.e. and) can be non-zero.

Multivariable Mendelian randomization (MVMR) can estimate the direct causal effect of multiple exposures, conditional on the other exposures.^17 18^ Thus, MVMR can estimate the direct effects of each educational level on an outcome, conditional on the other educational levels.^19^ MVMR, applied in this way, allows for the effect of the genetic variants on the educational levels to be accounted for in the estimation. MVMR relies on the same assumptions as univariable Mendelian randomization (relevance, independence, and exclusion), with the additional requirement that the instruments explain sufficient variation in each exposure conditional on the other exposures (conditional relevance). This requires the genetic instruments to have sufficient differential effects on each educational level to identify the effects on each period. Note that genetic instruments can affect all educational levels; there merely needs to be sufficient variation in the size of these effects. Conditional relevance can be tested with a conditional F-statistic.^20 21^

In this paper, we demonstrate how MVMR can estimate the direct effects of educational levels on later health outcomes. We use educational attainment data from UK Biobank^22^ and summary-data two-sample Mendelian randomization to efficiently estimate the direct causal effect of remaining in school to age 18 or achieving a university degree, compared to leaving school before age 18, on body mass index (BMI), smoking initiation, and systolic blood pressure. Our results demonstrate that we can identify the differential effects of educational levels and that for two of the outcomes considered (BMI and smoking initiation), there may be differences in the effects of achieving different levels of education.

## Methods

### Data

UK Biobank is a population-based health research resource consisting of 503,317 people aged between 38 years and 73 years who were recruited between 2006 and 2010 from across the UK. Participants reported their demographics, health status, lifestyle measures, cognitive testing, personality self-report, and physical and mental health measures) via questionnaires and interviews. A full description of the study design, participants and quality control (QC) methods have been described in detail.^22-24^ UK Biobank received ethics approval from the Research Ethics Committee (REC reference for UK Biobank is 11/NW/0382).

DNA was extracted from the genotyped blood samples using UK BiLEVE Axiom and UK Biobank Axiom arrays. Full details of the genotyping, imputation and quality control procedure are available elsewhere.^25 26^

Participants reported their educational qualifications. We defined each participant’s educational attainment as the highest qualification they reported. We then categorised these educational qualifications into three levels; left school around 16 (highest qualifications: GCSE’s/O-levels/CSEs or lower), completed school to age 18 (highest qualifications: A-levels) and has a college or university degree. Other qualifications were grouped based on the closest equivalent (see **Supplementary Table 1**).

#### Genome-wide Association Study of educational attainment

We ran an independent Genome-Wide association study (GWAS) to estimate the association of genetic variants with each of two binary variables: 1) remaining in school to at least 18 vs leaving before 18, and 2) having a university or college degree vs not.

Participants were coded 1/0 based on whether they had achieved a qualification associated with that educational level (or higher). The GWAS was conducted using BOLT-LMM,^27^ which controls for relatedness and population stratification and relatedness. We applied standard exclusions to the UK Biobank data, limiting our sample to participants of European ancestry, excluding participants who had withdrawn consent, and for other reasons.^26^ We further limited our sample to participants who reported being born in England to restrict to individuals who experienced similar education systems. We also adjusted for age and sex.

We identified genome-wide significant SNPs, defined as p<5×10^−8^ and independent (LD=10,000kb, r^2^ <0.01). These criteria were applied to each of the two GWAS results for the binary variables for the educational stage to select SNPs associated with that stage. To create a list of ‘hits’ associated with at least one educational level, we combined the hits selected from both GWAS and applied a further round of clumping to remove overlapping loci using the same parameters.

### Mendelian Randomization

We extracted the SNP-outcome associations for BMI, smoking initiation and SBP from a separate GWAS based on the largest GWAS studies available in OpenGWAS for each SNP identified as associated with educational attainment in our GWAS.^28 29^ We did not use GWAS that included only UK Biobank to minimise winner’s curse bias.^30^ We used a BMI GWAS of 681,275 individuals of primarily European ancestry, in which BMI was reported as a standard deviation change. 456,426 individuals were from the UK Biobank, representing about 67% of the sample.^31^ We used a smoking initiation GWAS of 607,291 individuals; 383,631 were from the UK Biobank, representing about 63% of the sample.^32^ This GWAS used a binary measure of smoking initiation defined as ever having smoked and reported the log odds of smoking from a logistic model. We used a systolic blood pressure (SBP) GWAS of 757,601 individuals of European ancestry; 458,577 were from the UK biobank, representing about 60% of the sample.^33^ In this GWAS, systolic blood pressure was adjusted for medication use by adding 15 mmHg for individuals reporting blood pressure lowing medication. We harmonised the exposure and outcome summary data and excluded palindromic SNPs with a minor allele frequency >0.4.

We used univariable Mendelian randomization to estimate the effect of each level of education, compared to achieving a lower level of education, on each outcome using inverse variance weighting(IVW).^34^ This analysis used genome-wide significant SNPs from the GWAS of each educational level as instruments. We investigated how robust our results are to violations of the third instrumental variable assumption using weighted median^35^, weighted mode^36^ and MR Egger^37^ estimators. Consistent results across these estimators suggest that findings are less likely to be driven by pleiotropic or heterogeneous effects. The effect estimates from the univariable Mendelian randomization can be interpreted as the effect of remaining in school to at least 18 compared to leaving school before age 18 and the effect of achieving a university degree compared to not. These point estimates require a fourth point identifying assumption, such as a constant effect of treatment or no simultaneous heterogeneity (NOSH), both of which identify the average treatment effect or a monotonic effect of the SNPs on the likelihood of remaining in school, which would identify the local average effects of staying in school (i.e. the effects of staying in education to each level for those whose choice was affected by the genetic).^14 38 39^

### Multivariable Mendelian Randomization

We used two-sample MVMR to simultaneously estimate the direct effects of staying in school to 18 and obtaining a university degree on each outcome. We included all SNPs associated with either level of educational attainment at genome-wide significance as instruments in this analysis. The estimates from this analysis identify the direct effect of remaining in school to age 18 but not going to university, compared to leaving school before 18 and the direct effect of getting a university degree compared to leaving school at age 18 or earlier. We estimated MVMR using inverse variance weighting and calculated conditional F-statistics and Q-statistics to test for heterogeneity.^17^

A fundamental assumption of MVMR is that the instruments are associated with each exposure strongly, given the other exposures included in the estimation. Violation of this assumption will decrease the precision of the results obtained and can bias the effect estimates. In summary-data MVMR, this assumption can be tested with a conditional F-statistic.^21^ The critical values to test for weak instrument bias are generally approximated to a ‘rule-of-thumb’ of 10; if the conditional F-statistics are larger than ten, then the SNPs are typically considered strong instruments and are unlikely to suffer from weak instrument bias.^40 41^

Patients and the public were not involved in designing the study.

## Results

### GWAS

We identified 360 independent SNPs associated with one of the measures of educational attainment at genome-wide significance levels (p < 5×10^−8^) (**Table 1**). Of these, 130 SNPs were significantly associated with leaving school at 18 and achieving a degree. However, a substantial proportion of SNPs were only associated at genome-wide significance levels with one or other education level, illustrated in **Figure 2**.

**Table 1.** Number of SNPs by educational level.

| Education level | No. SNPs |
| --- | --- |
| Remaining in school to 18 | 264 |
| Getting a degree | 226 |
| **Total** | **360** |

**Figure 2.**
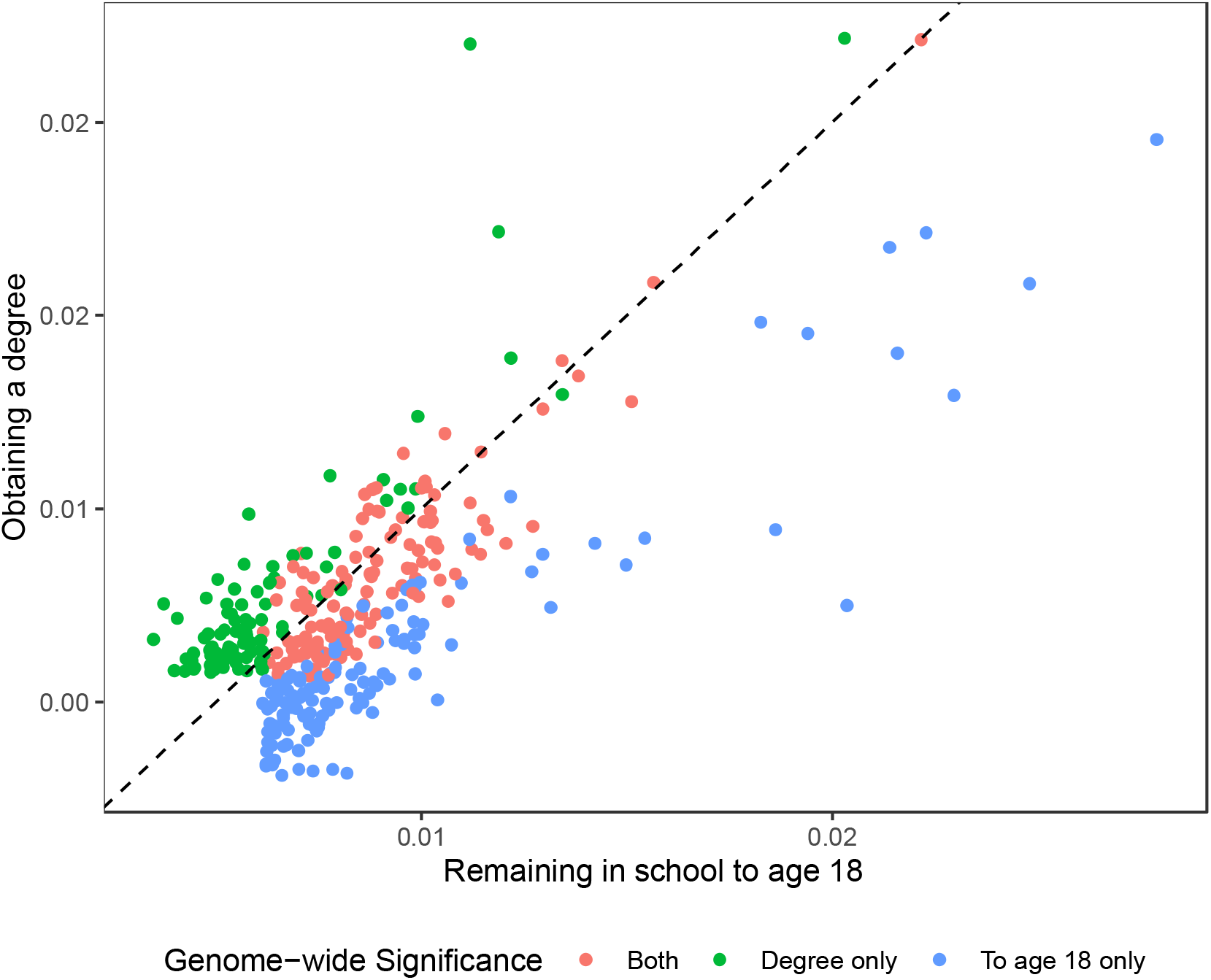
Association of SNPs and remaining in school after 18 and/or getting a degree. A plot of the association between each SNP and remaining in school to age 18 and obtaining a degree for all SNPs associated with either staying in school after 18 or having a degree at genome-wide significance. Colours show whether each SNP was associated with staying in school to 18, getting a degree, or both.

### Instrument strength testing

We tested whether the selected SNPs were sufficiently strongly conditionally associated with educational attainment using conditional F-statistics (**Table 2**). They lie between 2.64 and 2.86, indicating that our MVMR estimation may suffer from weak instrument bias. In MVMR, the direction of that bias can be either towards or away from the null, and therefore we cannot draw any conclusions about the impact of this bias. This reduction in the F-statistic between the univariable and MVMR is likely because of the high level of correlation between the effect of the SNPs on each measure of educational attainment, which will lower the power and instrument strength for MVMR.

**Table 2.** F-statistics for each level of educational attainment.

|  | **BMI** |  | **Smoking initiation** |  | **SBP** |  |
| --- | --- | --- | --- | --- | --- | --- |
|  | *F-statistic* | *Conditional F-statistic* | *F-statistic* | *Conditional F-statistic* | *F-statistic* | *Conditional F-statistic* |
| **Remaining in school after 18** | 42.21 | 2.65 | 42.01 | 2.70 | 42.05 | 2.64 |
| **Having a degree** | 43.03 | 2.75 | 44.09 | 2.86 | 44.09 | 2.86 |

Number of SNPs; BMI 254, smoking initiation 326, SBP 323. Phenotypic correlations between each educational attainment level calculated from individual-level data in UK Biobank. The number of SNPs in each estimation varies due to the availability of SNPs in the outcome data.

### Mendelian randomization results

The univariable and MVMR estimates of the effect of remaining in school to 18 and getting a degree are shown in **Figure 3 and Table 3**. Full results, including sensitivity analyses allowing for alternative structures of pleiotropy, are given in **Supplementary Table 3**. The univariable Mendelian randomization estimates suggested that remaining in school to at least 18 and getting a university degree had similar effects on BMI. Remaining in school to at least age 18 decreased BMI by 0.36 standard deviations, 95% confidence interval (95%CI): [0.25 to 0.47]. Getting a degree decreased BMI by 0.38 standard deviations, 95%CI: [0.25 to 0.52]. However, when both levels of educational attainment are included using MVMR, the effect of remaining in school to 18 attenuated (0.04, 95% CI: -0.42, 0.50). Very little change was seen in the point estimate for achieving a degree between the univariable MR and MVMR (MVMR estimated effect of having a degree; 0.47, 95% CI: [0.01 to 0.97].

**Table 3.** Univariable and multivariable MR estimates of the effect of remaining in school to 18 and having a degree on BMI, smoking initiation and systolic blood pressure.

|  | No.<br>SNPs | Effect<br>est. | 95 % C.I. | Q-statistic<br>p-value |
| --- | --- | --- | --- | --- |
| **BMI** |  |  |  |  |
| *Remaining in school to 18* |  |  |  |  |
| UVMR | 196 | -0.45 | [-0.56 -0.35] | 1.92x10<sup>-311</sup> |
| MVMR | 254 | 0.09 | [-0.34 0.51] | 2.26x10<sup>-321</sup> |
| *Getting a degree* |  |  |  |  |
| UVMR | 178 | -0.48 | [-0.60 -0.36] | 5.39x10<sup>-281</sup> |
| MVMR | 254 | -0.63 | [-1.10 -0.17] | 2.26x10<sup>-321</sup> |
| **Smoking Initiation** |  |  |  |  |
| *Remaining in school to 18* |  |  |  |  |
| UVMR | 258 | 0.53 | [0.47 0.59] | 2.15x10<sup>-78</sup> |
| MVMR | 326 | 0.48 | [0.30 0.76] | 3.35x10<sup>-87</sup> |
| *Getting a degree* |  |  |  |  |
| UVMR | 224 | 0.58 | [0.50 0.66] | 2.26x10<sup>-83</sup> |
| MVMR | 326 | 1.14 | [0.69 1.88] | 3.35x10<sup>-87</sup> |
| **SBP** |  |  |  |  |
| *Remaining in school to 18* |  |  |  |  |
| UVMR | 256 | -2.60 | [-3.73 -1.46] | 5.66x10<sup>-135</sup> |
| MVMR | 323 | -0.63 | [-5.33 4.06] | 6.50x10<sup>-170</sup> |
| *Getting a degree* |  |  |  |  |
| UVMR | 221 | -3.63 | [-4.92 -2.34] | 1.83x10<sup>-117</sup> |
| MVMR | 323 | -2.55 | [-7.73 2.63] | 6.50x10<sup>-170</sup> |

Mendelian randomization and MVMR effect estimates for remaining in school after 18 and having a degree estimated by Univariable Mendelian Randomization and Multivariable Mendelian Randomization. UVMR=inverse variance weighted univariable Mendelian randomization, MVMR=inverse variance weighted multivariable Mendelian randomization. BMI=Body mass index. SBP=systolic blood pressure. Effect estimates are standard deviation change in BMI, odds ratio for smoking initiation and mmHg unit change per unit change in remaining in school to 18/obtaining a degree. The number of SNPs varies across the analyses as not all SNPs were available in every outcome GWAS. Q statistic p-value is the p-value for a test of heterogeneity in the effect estimates across all SNPs. These are the same for each level of education in the MVMR analyses, as one statistic is reported per analysis.

**Figure 3.**
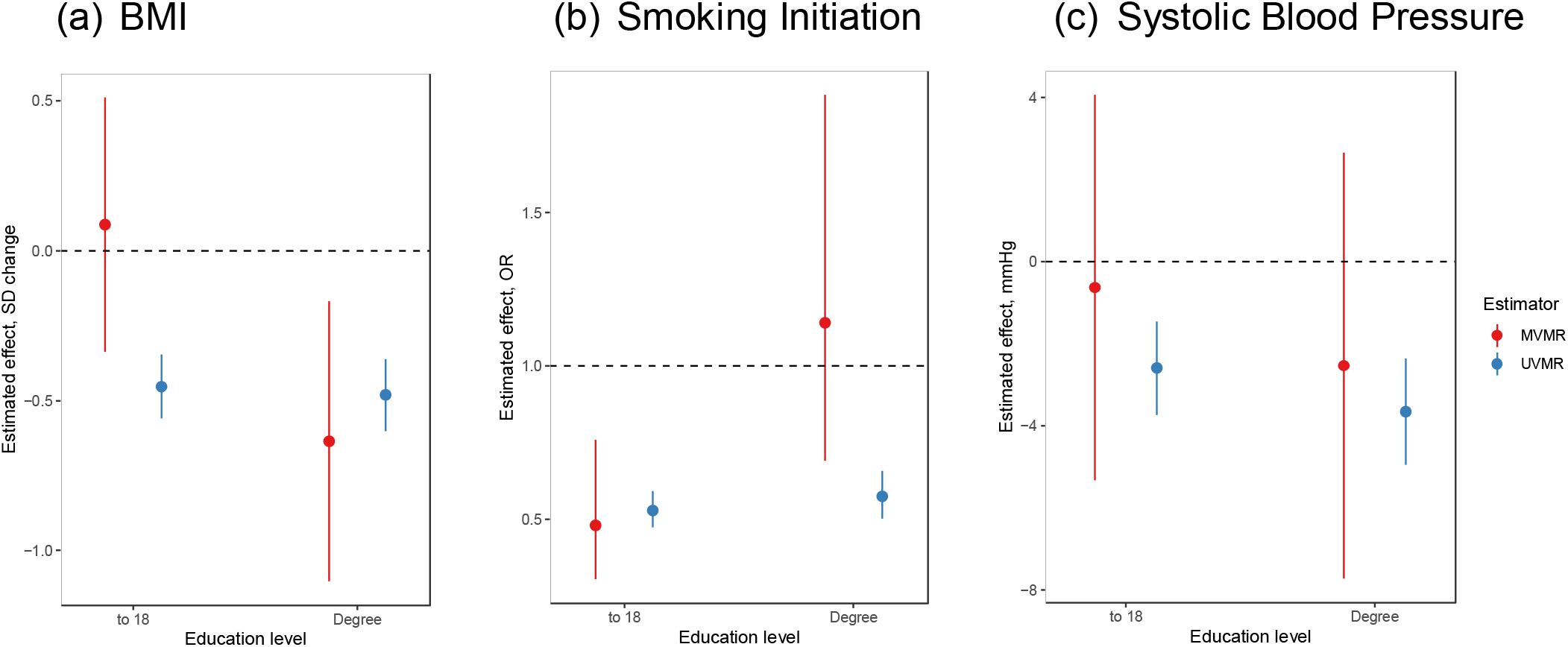
Univariable MR (blue) and MVMR (red) estimates for the effect of remaining in school to 18 and getting a degree on BMI, smoking initiation and systolic blood pressure. IVW and MVMR estimate effects of remaining in school after 18 and having a degree on BMI (standard deviation change), smoking initiation (odds ratio) and systolic blood pressure (change in mmHg). UVMR=Inverse variance weighted univariable Mendelian randomization, MVMR= inverse variance weighted multivariable Mendelian randomization. BMI=Body mass index. SBP=systolic blood pressure.

The univariable Mendelian randomization estimates suggested that remaining in school to at least 18 and obtaining a degree affected the risk of smoking initiation. Staying in school to 18 decreased the odds of smoking initiation (OR: 0.53, 95%CI: 0.47, 0.59), getting a degree reduced the odds of smoking initiation by a similar amount (OR: 0.58, 95%CI: 0.54 to 0.66). The MVMR estimates for remaining in school to 18 were consistent with the univariable estimates (OR: 0.48, 95% CI: 0.30 to 0.76). However, the MVMR estimates of the effect of getting a degree were entirely attenuated (OR: 1.14, 95% CI: 0.69 to 1.88).

The univariable Mendelian randomization estimates for SBP suggested that both remaining in school and having a degree reduced blood pressure. Remaining in school to at least 18 decreased SBP by 2.60mmHg (95%CI:1.46 to 3.73), and obtaining a university degree decreased SBP by 3.63mmHg (95%CI: 2.34 to 4.92). The MVMR estimates for both stages were attenuated towards the null; however these estimates were very uncertain and were also consistent with the univariable analysis.

## Discussion

MVMR can estimate the effect of time-varying exposures on later outcomes. Here we demonstrated that MVMR can be used to estimate the effects of levels of educational attainment on a range of health outcomes. Previous studies have assumed that educational attainment is a continuous variable scaled by years of education. This simplifying assumption increases statistical power. However, it limits the policy relevance of the estimates. The effects of remaining in school until age 18 likely differ from those of getting a university degree. For example, our MVMR estimates suggest that staying in school until age 18 lowers the rate of smoking initiation but that achieving a degree has little additional benefit. In contrast, our MVMR results suggest that remaining in school until age 18 has little effect on BMI but that getting a university degree reduces BMI. This approach provides valuable evidence about when educational effects occur, which can give clues on aetiology and potential intervention strategies and the external validity of Mendelian randomization estimates.

MVMR estimates are the direct effect of achieving each educational level, conditional on the other levels of educational attainment. We used the lowest level of educational attainment as our reference category, so the estimated effect of remaining in school to age 18 but not getting a degree, compared to leaving at age 16 or before. The estimated effect is getting a degree is compared to leaving school at age 18. In contrast, the univariable Mendelian randomization estimates of the effect of a degree reflect the effect of getting a degree versus not getting a degree. Furthermore, the MVMR estimates have less restrictive assumptions about the structure of pleiotropy. MVMR requires the assumption that there are no pleiotropic effects of variants which affect the outcome, which is not mediated via either of our measures of educational attainment. The effects of remaining in school to 18 will reflect many potential pleiotropic pathways (e.g., baseline ability). However, these pathways are unlikely to bias the estimated effect of getting a university degree in the MVMR estimation, as they have been accounted for through the exposure indicating remaining in school to 18. So, the results obtained are unlikely to be explained by the pleiotropic effects via ability.

In general, MVMR estimators are generally less precise than univariable MR estimators. This is because MVMR only uses conditional variation in the exposure, which explains less of the overall variation in the exposures. This reduces the instruments’ strength and the analyses’ statistical power. As a result, MVMR requires very large sample sizes to estimate the effects of each level of educational attainment precisely. A limitation of our analysis is the conditionally weak instruments seen in our MVMR, and correspondingly large confidence intervals. Weak instruments can result in imprecise estimates. However, we had sufficient power to detect education stage specific effects on BMI and smoking initiation. Here, we only used a relatively small sample size of 371,053 to estimate the SNP-education associations. Future studies should run GWAS on binary outcomes of these educational stages to provide instruments to investigate the effects of these critical educational levels. This would allow more precise estimates of the effects of each educational level and provide more evidence about how, why and when educational attainment affects health outcomes.

A further limitation that applies to all Mendelian randomization studies is that selection bias may induce bias in our results. This could occur if both educational attainment and our outcomes influence selection into our study. The participants of the UK Biobank were far more educated and healthy than the general population.^24^ Future studies should use inverse probability weighting to account for the study’s sampling. Our results may reflect the pleiotropic effects of the SNPs, which directly affect the outcome. However, these pleiotropic effects would have to be independent of remaining in school to 18 and thus likely exclude many common explanations (e.g., ability). We also investigated the level of heterogeneity in our results and used alternative estimators which make different assumptions about the structure of pleiotropy. The alternative univariable Mendelian randomization estimators gave results consistent with the IVW effect estimates. The lower precision in MR Egger and weighted Mode estimates is expected as these estimators have lower power than IVW and weighted Median. However, our estimates showed high levels of heterogeneity across both exposures highlighting the potential for further pleiotropic effects. Finally, instrumental variable estimates require a fourth point identifying assumption. Under a constant effect of education or no simultaneous heterogeneity (NOSH), our results will reflect the average treatment effect. Under monotonicity, our results will reflect the local average treatment effects (i.e., the effects of remaining in school to age 18 or university for those whose decision was affected by these variants).

In summary, we can use MVMR to estimate time-varying effects, such as the effects of different educational levels. We can improve the power and precision of these estimates by efficiently combining studies and could potentially provide new insights into when and how educational attainment affects health outcomes.

## Data Availability

The UK Biobank data used in this study is available on application to UK Biobank. All other data is available online at https://gwas.mrcieu.ac.uk/.

## Contributions

ES designed the study, wrote the first draft, revised the manuscript, and analysed the data. NMD designed the study, revised the manuscript and analysed the data.

## Data availability

The individual data used for the GWAS study was analysed under UK Biobank application number 81499. The analytic datasets used in the study have been [reviewer note - will be] archived with the UK Biobank study. Please get in touch with for further information.

All other analysis was conducted using publicly available data accessed through the IEU OpenGWAS infrastructure.

## Code availability

All analyses were conducted in R version 4.2.2, The code used to generate these results has been archived at (https://github.com/eleanorsanderson/EducationMVMR).

