## Supplementary Table 1 for "The relative effects of staying in school and attending university on smoking, BMI and systolic blood pressure: evidence from multivariable Mendelian randomization"

**SUPPLEMENTARY MATERIAL**

Eleanor Sanderson^1,2^ and Neil M. Davies^1,3,4,5^

1. Medical Research Council Integrative Epidemiology Unit, University of Bristol, BS8 2BN, United Kingdom.
2. Population Health Sciences, University of Bristol, Barley House, Oakfield Grove, Bristol, BS8 2BN, United Kingdom.
3. Division of Psychiatry, University College London, Maple House, 149 Tottenham Court Rd, London W1T 7NF
4. Department of Statistical Sciences, University College London, London WC1E 6BT, UK
5. K.G. Jebsen Center for Genetic Epidemiology, Department of Public Health and Nursing, Norwegian University of Science and Technology, Norway.

***Supplementary Table 1 – Qualifications reported in UK Biobank and corresponding educational attainment level***

| *Qualification* | *Educational attainment level* | *% as highest qualification* |
| --- | --- | --- |
| College or university degree | Obtained a degree | 32.9% |
| Other professional qualifications (e.g. nursing, teaching) | Remained in school to 18 | 14.7% |
| A levels/AS levels or equivalent | Remained in school to 18 | 7.4% |
| NVQ, HND, HNC or equivalent | Left school before 18 | 5.9% |
| O levels/GCSEs or equivalent | Left school before 18 | 17.0% |
| CSEs or equivalent | Left school before 18 | 4.8% |
| None | Left school before 18 | 17.5% |

Categorisation of the different highest educational qualifications reported in UK Biobank. % as highest qualification gives the proportion of UK Biobank for who this qualification is the highest reported.

***Supplementary Table 2 – Sample sizes for the GWAS***

| ***Education level*** | ***n*** | ***No*** | ***Yes*** | ***% Yes*** |
| --- | --- | --- | --- | --- |
| **Remained in school to 18** | 370,953 | 173,751 | 197,202 | 53.2% |
| **Obtained a degree** | 370,861 | 256,654 | 114,207 | 30.8% |

Sample sizes for the genome-wide association studies conducted and proportion of individuals who had obtained at least that level of qualification in each one.

***Table 3 – Univariable and multivariable MR estimates of the effect of remaining in school to 18 and having a degree on BMI, smoking initiation and systolic blood pressure.***

|  |  | ***No. SNPs*** | ***Effect est.*** | ***95 % C.I.*** | | ***Q-statistic***  ***p-value*** |
| --- | --- | --- | --- | --- | --- | --- |
| ***BMI*** |  |  |  |  | |  |
|  | *Remaining in school to 18* | | |  |  |  |
|  | **IVW** | 196 | -0.45 | [-0.56 | -0.35] | 1.92x10^-311^ |
|  | **W.Median** | 196 | -0.37 | [-0.44 | -0.29] |  |
|  | **W.Mode** | 196 | -0.63 | [-1.00 | -0.25] |  |
|  | **MR Egger** | 196 | -0.18 | [-0.72 | 0.36] |  |
|  | **MVMR** | 254 | 0.09 | [-0.34 | 0.51] | 2.26x10^-321^ |
|  | *Getting a degree* | | |  |  |  |
|  | **IVW** | 178 | -0.48 | [-0.60 | -0.36] | 5.39x10^-281^ |
|  | **W.Median** | 178 | -0.37 | [-0.46 | -0.29] |  |
|  | **W.Mode** | 178 | -0.19 | [-0.55 | 0.17] |  |
|  | **MR Egger** | 178 | 0.04 | [-0.52 | 0.61] |  |
|  | **MVMR** | 254 | -0.63 | [-1.10 | -0.17] | 2.26x10^-321^ |
| ***Smoking Initiation*** | |  |  |  |  |  |
|  | *Remaining in school to 18* | | |  |  |  |
|  | **IVW** | 258 | 0.53 | [0.47 | 0.59] | 2.15x10^-78^ |
|  | **W.Median** | 258 | 0.56 | [0.51 | 0.63] |  |
|  | **W.Mode** | 258 | 0.46 | [0.34 | 0.64] |  |
|  | **MR Egger** | 258 | 0.66 | [0.41 | 1.07] |  |
|  | **MVMR** | 326 | 0.48 | [0.30 | 0.76] | 3.35x10^-87^ |
|  | *Getting a degree* | | |  |  |  |
|  | **IVW** | 224 | 0.58 | [0.50 | 0.66] | 2.26x10^-83^ |
|  | **W.Median** | 224 | 0.61 | [0.54 | 0.69] |  |
|  | **W.Mode** | 224 | 0.52 | [0.34 | 0.79] |  |
|  | **MR Egger** | 224 | 0.76 | [0.43 | 1.34] |  |
|  | **MVMR** | 326 | 1.14 | [0.69 | 1.88] | 3.35x10^-87^ |
| ***SBP*** |  |  |  |  |  |  |
|  | *Remaining in school to 18* | | | |  |  |
|  | **IVW** | 256 | -2.60 | [-3.73 | -1.46] | 5.66x10^-135^ |
|  | **W.Median** | 256 | -2.74 | [-3.68 | -1.81] |  |
|  | **W.Mode** | 256 | -1.72 | [-5.70 | 2.26] |  |
|  | **MR Egger** | 256 | -3.59 | [-8.73 | 1.55] |  |
|  | **MVMR** | 323 | -0.63 | [-5.33 | 4.06] | 6.50x10^-170^ |
|  | *Getting a degree* |  |  |  |  |  |
|  | **IVW** | 221 | -3.63 | [-4.92 | -2.34] | 1.83x10^-117^ |
|  | **W.Median** | 221 | -3.43 | [-4.53 | -2.33] |  |
|  | **W.Mode** | 221 | -5.54 | [-10.07 | -1.01] |  |
|  | **MR Egger** | 221 | -0.35 | [-6.01 | 5.31] |  |
|  | **MVMR** | 323 | -2.55 | [-7.73 | 2.63] | 6.50x10^-170^ |

Mendelian randomization and MVMR effect estimates for remaining in school after 18 and having a degree estimated by Inverse variance weighting (IVW), Weighted median (W.Median), Weighted mode (W.Mode), MR Egger and MVMR-IVW. MVMR=multivariable Mendelian randomization. BMI=Body mass index. SBP=systolic blood pressure. The number of SNPs varies across the analyses as not all SNPs were available in every outcome GWAS. Q statistic p-value is the p-value for a test of heterogeneity in the effect estimates across all SNPs. These are the same for each level of education in the MVMR analyses, as one statistic is reported per analysis.
